# Two viruses competition in the SIR model of epidemic spread: application to COVID-19

**DOI:** 10.1101/2022.01.11.22269046

**Authors:** A.M. Ignatov, S.A. Trigger

**Affiliations:** Prokhorov General Physics Institute of the Russian Academy of Sciences, 38 Vavilova St., Moscow, 119991 Russia; Joint Institute for High Temperatures, Russian Academy of Sciences, 13/19, Izhorskaia Str., Moscow 125412, Russia

## Abstract

The SIR model of the epidemic spread is used for consideration the problem of the competition of two viruses having different contagiousness. It is shown how the more contagious strain replaces over time the less contagious one. In particular the results can be applied to the current situation when the omicron strain appeared in population affected by the delta strain.

**PACS number(s):** 02.50.-r, 05.60.-k, 82.39.-k, 87.19.Xx

---

The most of existing models for the spread of infection simulate the spontaneous development of an epidemic and describe all its stages. There are two kinds of such models: susceptible-infected-susceptible (SIS) models and susceptible–infectious–removed, susceptible–exposed–infectious–removed (SIR, SEIR) models. The first one goes back to the pioneering work of Kermack and McKendrick [1] and uses the assumption that the recovered people can immediately get infection again. On the contrary, the latter are built on the assumption that the recovered people save strong immunity during epidemic (see, e.g. [2]). There are many variants of those models.The SIS models are used in the mathematical epidemiology [3]. An overview is given in [4–6] (see also references therein). Balance between the susceptible and infected members of population under the various conditions of infection transfer, are the subject of research in [7–9].

Developing the delayed time-discrete epidemic model (DTDEM) [10] the papers [11, 12] take into account delay features of COVID-19 in differential form. An important feature of this model is the delay, which takes into account the long-term carriage of viruses confirmed by clinical data. In contrast to the delay discussed in [13], the discrete delay model assumes that the patient is immune, and in this respect fits the SIR model, not SIS. At the same time, the considered delay model does not imply the allocation of a separate category of hidden virus carriers (see, for example, the SEIR model with dealy in [14]). Latent carriers of the virus can infect others without delay and in this sense are similar to those infected, in contrast to the SEIR model. Currently, the simplest SIR and SEIR baseline models are being developed taking into account the vaccination process [15–17].

In this paper our purpose is to consider the SIR-type equations for circulation of two viruses in the finite population *N*. We suppose for simplicity that one person can only carry one virus and after disease the recovered people cannot be infected again by any of the viruses. These properties of the considered viruses we call for shortness “virus orthogonality” in conventional analogy with mathematical orthogonality of functions. Naturally, in contrast with mathematics both properties are restricted and reflect statistical observations. There is only limited period of immunity, however much longer than the average disease duration. At the same time disease COVID-19 caused by two stains which simultaneously present in one sick person is not known (in contrast with the rare cases of COVID-19 and flu).

The model can be easily extended to take into account vaccination and different quarantine measures. Below we consider the simple case of the free running epidemic under two viruses circulation. This assumption can be considered as realistic for very fast developing epidemic caused by, e.g., the omicron strain (or another highly contagious virus) appeared in a population affected earlier by a less contagious virus.

In general case, the SIR model (or SEIR, or DTDEM models) can be generalized for the case of two viruses circulation by inclusion of various quarantine measures, e.g., through the so-called function of quarantine measures influence, accounting the government restrictions, vaccination process etc. Also the cases of death, re-infection with one of the viruses some time after recovery, limited time for effective vaccination and other known factors should be included in a general model. However, below we consider the simplest SIR model to clarify the main specific features of competition of two viruses in population. The generalization considered below seems useful for any more advanced models describing epidemic processes with actively mutating viruses.

Let us introduce *S* - the number of never non-infected people in a population, *I*_1_ and *I*_2_ - the number of viruses carriers of type 1 and 2. Then equations of the generalized SIR model is written in the form

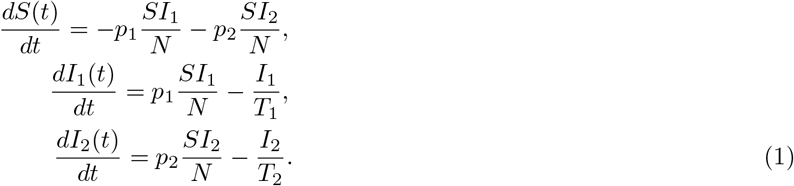

where the parameters *p*_1_ and *p*_2_ are the characteristics of the contagiousness for two viruses, *T*_1_ and *T*_2_ are the characteristic average time of the diseases caused by viruses of the types 1 and 2.

The evident equalities are: *I*(*t*) = *I*_1_(*t*) + *I*_2_(*t*) is the full numbers of virus carriers at the moment *t*, which is the sum of carriers *I*_1_ of the first and *I*_2_ the carriers of the second virus. Introduce also the full numbers of people infected by some of two viruses to the moment of time *t* (ill and recovered=affected by the viruses) is *N*_*tot*_(*t*) = *N*_1_(*t*) + *N*_2_(*t*), where *N*_*i*_(*t*) (i=1,2) is the number of people affected (ill and recovered) by virus of type *i* to the moment *t*.

Therefore *N* = *S*(*t*) + *N*_1_(*t*) + *N*_2_(*t*) ≡ *S* + *N*_*tot*_ = *Const*., where 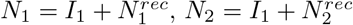 and 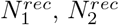-the full numbers of recovered after disease by the viruses 1 and 2, respectively.

Now rewrite equations (1) by use the variables *I*_1_(*t*)*/N* = *y*_1_(*t*), *I*_2_(*t*)*/N* = *y*_2_(*t*), *S/N* = *u*(*t*)

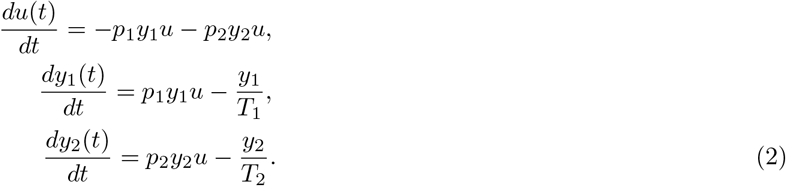

The value *u*(*t*) ≡ 1 − *z*(*t*) is the depletion of the population of non-affected by viruses people, therefore *z*(*t*) = *N*_*tot*_(*t*)*/N* corresponds to the fraction of full population *N* that are affected (ill and recovered) by both viruses to the time *t*. The values *y*_1_(*t*) and *y*_2_(*t*) are the current fractions of population actively infected by the viruses 1 and 2 (viruses carriers) in a moment *t*, respectively.

The stationary solution 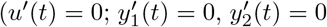, where stroke over the letter denotes the time derivative) to Eqs. (2) exists only if *y*_1_(*t*) = 0, *y*_2_(*t*) = 0 and *u*(*t*) = *U*, where *U* is an arbitrary constant. Linearization of Eqs. (2) near this solution gives the condition of stability

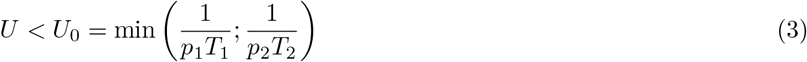

Corresponding solution Eq. (1) at *t* → ∞ is

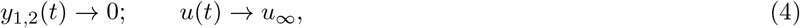

where *u*_∞_ is a certain constant, moreover *u*_∞_ < *U*_0_. It is the estimate for the saturation value.

The consequence of Eq. (1) is *u*^′^(*t*) < 0. If *u*(*t* = 0) = *u*_0_ and the value *p*_*i*_*u*_0_ < 1*/T*_*i*_ we conclude that 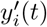 is a monotonously decreasing function, otherwise *y*_*i*_(*t*) should reach a maximal value.

Consider first as an example the case *p*_1_*u*_0_*T*_1_ > 1 and *p*_2_*u*_0_*T*_2_ < 1. In this case the second virus appeared in a small quantity in a population cannot actively spread even in the case *p*_2_ > *p*_1_ and function *y*_2_(*t*) monotonically decreases. The respective behavior is shown in Fig. 1, where the spread of only one virus is shown for the parameters *p*_1_ = 0, *p*_2_ = 0, *y*_1_(0) = 0.01, *y*_2_(0) = 0, *u*(0) = 1 − 0.01 and the average duration of the virus carrier *T*_1_ = 20 days.

**Figure 1:**
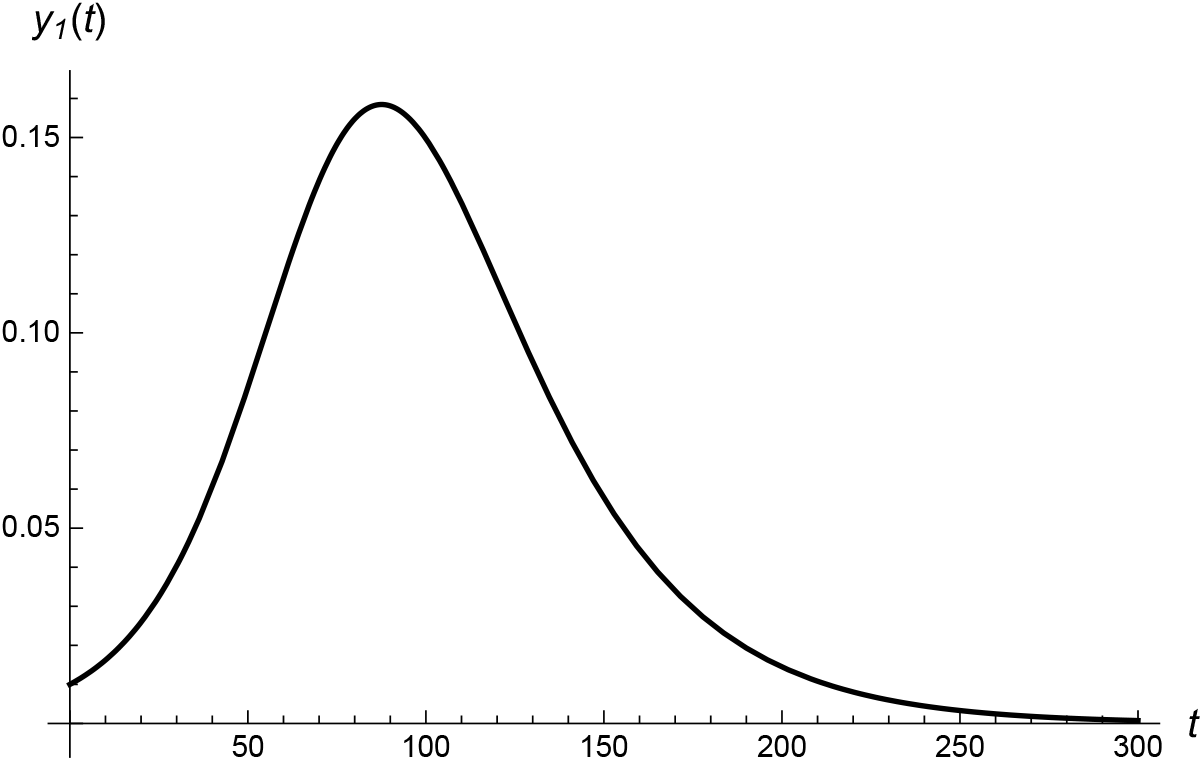
Function *y*_1_(*t*) of virus 1 carriers for the case when second virus is absent at all (time in days). The parameters are *p*_2_ = 0, *y*_1_(0) = 0.01, *u*(0) = 0.99 and the average duration of the virus carrier *T*_1_ = 20 days.

The most interesting case corresponds to the conditions *p*_*i*_*u*_0_*T*_*i*_ > 1, when both viruses spread infection by increasing the values *y*_*i*_(*t*). If at the initial stage of the epidemic the second virus is absent the first one spreads till some initial level *y*_1_(0). We consider t=0 as the moment of the second virus with higher contagiousness. We will suppose, according to the existing statistical data [18] that 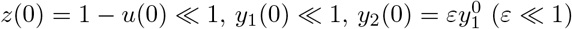. We also use the inequality *p*_1_ < *p*_2_ which means that the appeared in a small number of infected people virus 2 possesses more high than virus 1 contagiousness.

Let us consider as an typical example of two viruses circulation in a population the concrete initial conditions *y*_1_(0) = 0.01, *y*_2_(0) = 0.001 *u*(0) = 1 − 0.01 − 0.001. The parameters *p*_1_ = 0.1, *T*_1_ = 0.1 are the same as for calculations shown in Fig.1. The characteristic parameters for the second more contagious virus are *p*_2_ = 0.2, *T*_2_ = 20 days. The result of the calculations is drawn in Fig.2.

As is easy to see that virus 1 is effectively suppressed by virus 2 since the value of the maximum for the solid curve in Fig.2 more than three times lower than in Fig.1. Comparison of these figures shows that duration of virus 1 circulation is also effectively suppressed due to appearance of virus 2. It is easy to see also that the maximum for virus 1 in Fig. 2 is shifted to earlier time in comparison with Fig. 1. It follows from the relations *p*_*i*_*T*_*i*_*u*(*t*_*i,max*_) = 1 for maximums of functions *y*_*i*_(*t*) and the condition *u*^′^(*t*) < 0. Denote 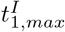 and 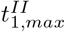 the times of maximal current of virus 1 carriers on Fig. 1 (case *I*) and Fig. 2 (case *II*) respectively. The first one is determined by the equality 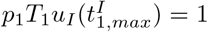, the second one by the equality 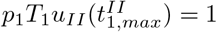, where the functions *u*_*I*_ and *u*_*II*_ correspond to the cases *I* and *II* respectively. Taking into account that *u*^′^(*t*) < 0 and according to the first equation (1) 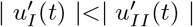 we arrive at the explicit inequality 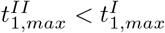.

**Figure 2:**
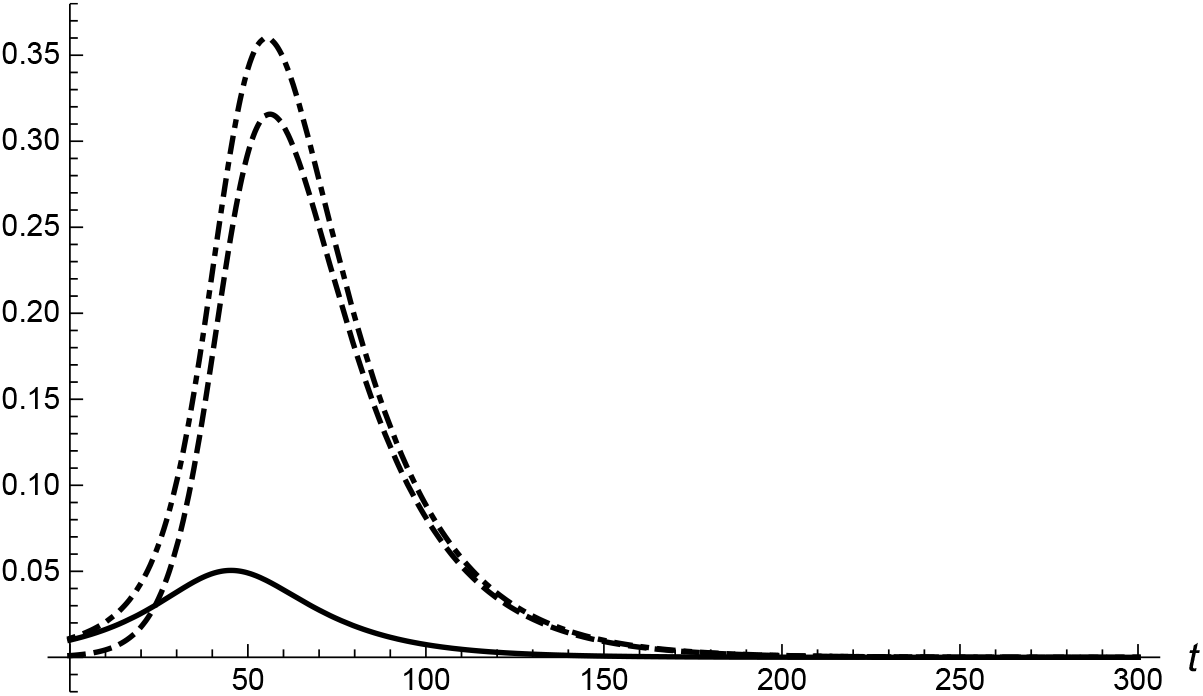
Fractions of the viruses 1 and 2 carriers as a function of time (in days): *y*_1_(*t*) for virus 1 (solid line), *y*_2_(*t*) for more contagious virus 2 (dashed line) and the sum *y*_1_(*t*) + *y*_2_(*t*) (dashed-dotted line) of the infected at moment *t*. The parameters *p*_1_ = 0.1, *p*_2_ = 0.2, *T*_1_ = *T*_2_ = 20 days, *u*(0) = 0.989.

Time behavior of function *u*(*t*) for the cases *I* and *II* is depicted in Fig.3. We see the more fast flowing epidemic and more full depletion of the set of healthy people.

It is also essential to find the fractions of people affected (ill and recovered) by the viruses of type 1 and 2 respectively to the moment *t* denoted as *x*_1_(*t*) and *x*_2_(*t*). To find *x*_1_(*t*) and *x*_2_(*t*) we use the relations

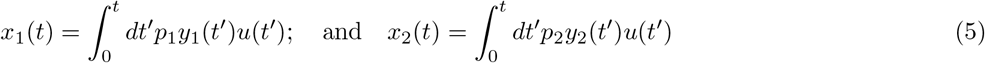

The result of calculation of functions *x*_1_(*t*) and *x*_2_(*t*) is presented in Fig. 4. Saturation of the epidemic in the case *II* takes place on the essential higher level (for the used parameters the full quantity of the affected by virus 2 people exceeds ones by virus 1 more than in four times).

**Figure 3:**
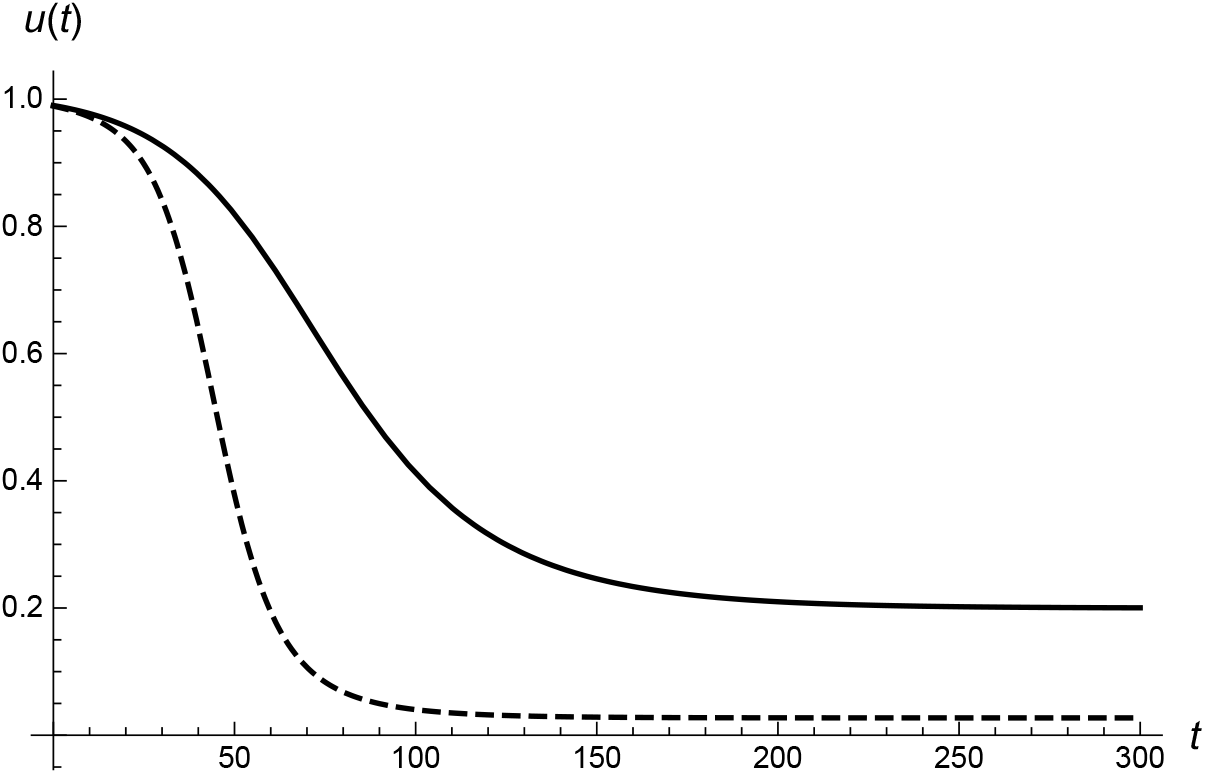
Time behavior of function *u*(*t*) for the cases *I* and *II*.

**Figure 4:**
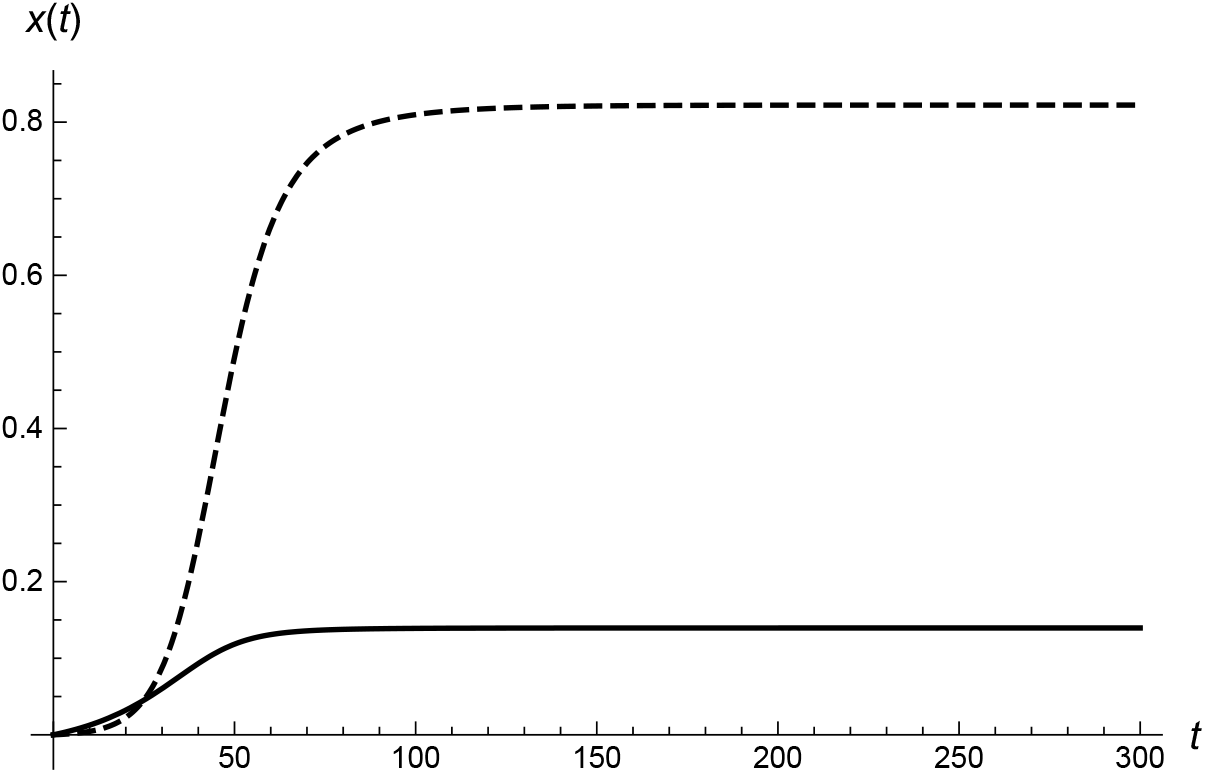
The functions *x*1(*t*) and *x*2(*t*) of the affected by the viruses 1 and 2 people in the case *II*.

The main results of the performed consideration for the SIR model generalized for free-running epidemic of two “orthogonal” viruses show the rapid displacement of a less infectious virus from a population by more contagious strain. The maximum value of the initially existed strain shifts to an earlier time and essentially decreases. The epidemic is progressing much faster and affects many more people. The inclusion of the effective quarantine measures can change the epidemic spread. However, these measures can be essentially restricted if the disease caused by the second strain is, on average, much easier. The obtained results are in general in an a good agreement with the qualitative discussions of experts in epidemiology and virology. In the connection of the obtained results the problem of the artificial creation of the “orthogonal” strains of the viruses SARS or some other “orthogonal” viruses, which causes the milder disease, can be discussed in future.

## Data Availability

All data produced in the present work are contained in the manuscript

## Acknowledgment

S.T. is thankful to infectious disease physician Dr. M. Karavaeva, Prof. Dr. F. Onufrieva and colleagues from the clinic Charité (Berlin) for many useful discussions.

